# Task-Adaptive Wasserstein Barycentric Fusion for Multimodal Medical Imaging

**DOI:** 10.64898/2026.04.05.26350202

**Authors:** Peijie Qiu, Zhaoqi An, Sungmin Ha, Sayantan Kumar, Aristeidis Sotiras

**Author notes:** (A. Sotiras).

## Abstract

Multimodal medical image analysis requires fusion that accounts for task-dependent modality contributions while preserving shared and modality-specific information. To address these needs, we propose the Task-Adaptive Wasserstein Barycenter Variational Autoencoder (*Wℬ*VAE-TA). The model forms a shared latent distribution as a Wasserstein barycenter of modality-specific encoding distributions. To adapt fusion to the task, we learn the modality weights jointly with the application objective while retaining closed-form aggregation of diagonal Gaussian distributions. To model modality-specific characteristics alongside the fused shared representations, the model uses multilevel decoding with modality-specific decoders and learnable modality embeddings. We evaluate *Wℬ*VAE-TA in brain tumor segmentation with complete and incomplete MRI inputs and in normative modeling with complete structural and diffusion-derived measurements. On BraTS 2018, the model achieves mean Dice scores of 62.31%, 78.83%, and 87.30% for enhancing tumor, tumor core, and whole tumor, respectively, averaged across all 15 modality combinations. Ablation studies demonstrate consistent gains from adaptive fusion and modality-specific conditioning. In normative modeling, the model achieves the highest estimated data log-likelihood among the evaluated methods on UK Biobank and ADNI, together with the highest ADNI significance ratio, precision, and balanced accuracy. These results support task-adaptive barycentric fusion with modality-specific conditioning across discriminative and generative medical imaging tasks.

## 1. Introduction

Multimodal learning is increasingly central to medical image analysis, where different imaging modalities provide complementary observations of anatomy and disease [1– 3]. In brain tumor analysis, multi-contrast Magnetic Resonance Imaging (MRI) sequences, including T1-weighted (T1w), T1 contrast-enhanced (T1ce), T2-weighted (T2w), and Fluid Attenuation Inversion Recovery (FLAIR), provide complementary cues for delineating tumor subregions [1, 4]. In normative modeling, structural and diffusion-derived measurements describe different aspects of brain morphology and microstructure, allowing individual variation to be examined across modalities [5]. A central challenge across these applications is to learn a representation that captures cross-modal relationships while retaining the distinctive information supplied by each modality.

Deep generative models, in particular variational autoen-coders (VAEs) [6], offer a principled route for this purpose. Prior studies have examined which VAE latent factors influence reconstruction and downstream prediction [7], and how global dependencies among observations can be learned without supervision [8]. In multimodal settings, a VAE combines unimodal encoding distributions into a joint latent distribution and decodes modality-specific likelihoods to reconstruct the observations [9, 10]. This generative objective encourages the learned representation to explain information across modalities and supports learning from multi-view observations without requiring disease labels.

However, *how to fuse the information across different modalities to learn faithful multimodal distributions* remains a central question. Most prior work approaches this problem primarily from a probabilistic view. Product-of-Experts (PoE) [9] and Mixture-of-Experts (MoE) [10] are two widely adopted probabilistic models for approximating the joint multimodal latent distribution. Subsequent variants, such as MoPoE [11], further build on and combine these two paradigms. Although empirical studies have demonstrated their success in multimodal VAEs, the theoretical understanding of their behavior remains underexplored [12]. For instance, PoE has been observed to bias the joint distribution toward certain modalities, whereas MoE can yield more coherent joint generation but may sacrifice sharpness or discriminability [11–13]. This is known as the bias-variance trade-off in mass allocation [12]. These observations can be traced to the underlying geometry implicit in these probabilistic models, particularly *how probability mass is allocated across modalities*.

To understand the properties of multimodal mass allocation, our earlier work [12] advocated a geometric view of multimodal VAEs, which formulated multimodal fusion as barycenter estimation and introduced Wasserstein Barycenter VAE (*Wℬ*VAE). Within this framework, a barycenter minimizes a weighted sum of squared 2-Wasserstein distances to the unimodal VAE encoding distributions. The resulting barycentric distribution serves as a fused approximate joint latent posterior conditioned on the available multimodal observations. Sampling from this posterior can then be used to reconstruct modality-specific observations through the VAE decoders. This geometric view exposes a trade-off between flexibility in mass allocation and tractability of fusion. The *αβ* divergence family allows its parameters to change how probability mass is allocated across modality distributions, but its barycenters generally lack closed-form solutions, including for Gaussian inputs. Adaptive *Wℬ* fusion offers an alternative route: for diagonal Gaussian encoders, modality weights adjust the fused location and spread while retaining closed-form aggregation. The illustration in Fig. 1 shows that suitable weights can approximate selected mass-allocation behaviors of numerical *αβ* barycenters, with residual shape differences. Adaptive Gaussian Wasserstein fusion interpolates the component means and standard deviations; it cannot produce a standard deviation below the smallest component value or represent multimodal densities. This motivates learning the weights directly through the medical imaging task objective, rather than numerically estimating an *αβ* barycenter within each fusion step.

**Figure 1.**
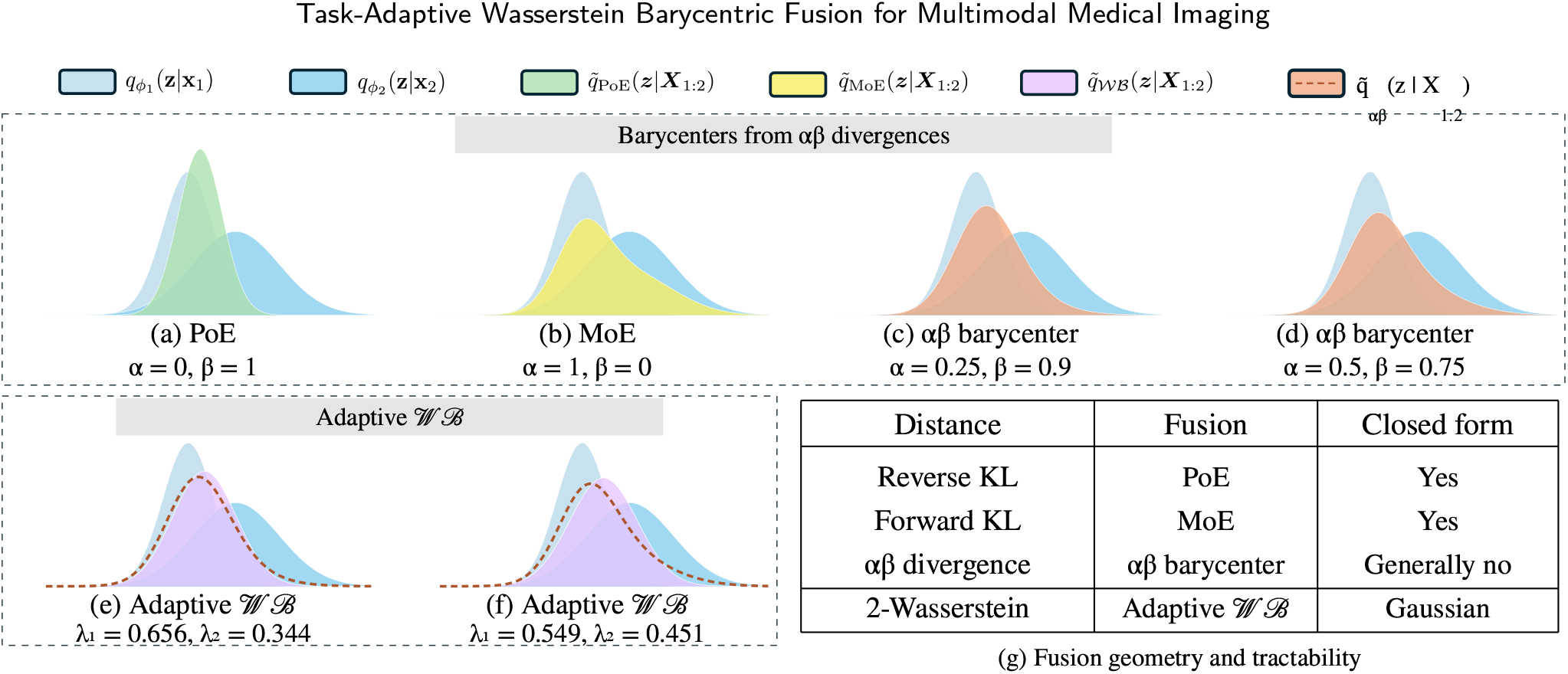
Illustration of multimodal Gaussian fusion. Panel (a) shows the standard PoE, whose Gaussian precision is the sum of component precisions; it is a reference product rather than the normalized-weight reverse-KL barycenter. Panels (b)–(d) compare barycenters associated with the *αβ* divergence family, including MoE as a special case. Panels (e)–(f) show Adaptive *Wℬ* approximations to the numerical targets in (c)–(d), with target-fitted weights and dashed target curves. The weights in (e)–(f) are fitted by minimizing the squared 2-Wasserstein distance to the corresponding numerical targets. Panel (g) summarizes fusion geometry and tractability.

However, the prior multimodal VAE formulations summarized in Table 1, including PoE-, MoE-, and MoPoE-based VAEs and *Wℬ*VAE, center representation learning on a shared fused posterior without an explicit modality-specific conditioning pathway across network levels. This emphasis is consistent with applications involving natural images and text, where the two modalities often convey shared semantic content: an image and its accompanying description can represent the same objects, attributes, or events. In contrast, medical imaging applications motivate two complementary requirements beyond shared fusion. First, *shared representations should be complemented by modality-specific modeling capacity*. In multimodal MRI, shared anatomy provides a consistent context for integrating evidence and localizing target regions, while contrast-specific appearances supply complementary cues for delineation. For example, T1ce highlights enhancing tumor, whereas FLAIR provides cues about peritumoral edema [1]. Similarly, structural and diffusion-derived measurements capture related but distinct aspects of brain morphology and microstructure [5]. Emphasizing cross-modal agreement alone may overlook these informative differences, motivating a shared–specific architecture in which shared latent representations are accompanied by an explicit modality-dependent decoding pathway. Second, *the relative contribution of each modality should adapt to the application*. Fusion weights suitable for brain tumor segmentation may not be appropriate for characterizing healthy variation in normative modeling. This motivates learning global fusion weights during task-specific training.

**Table 1.**
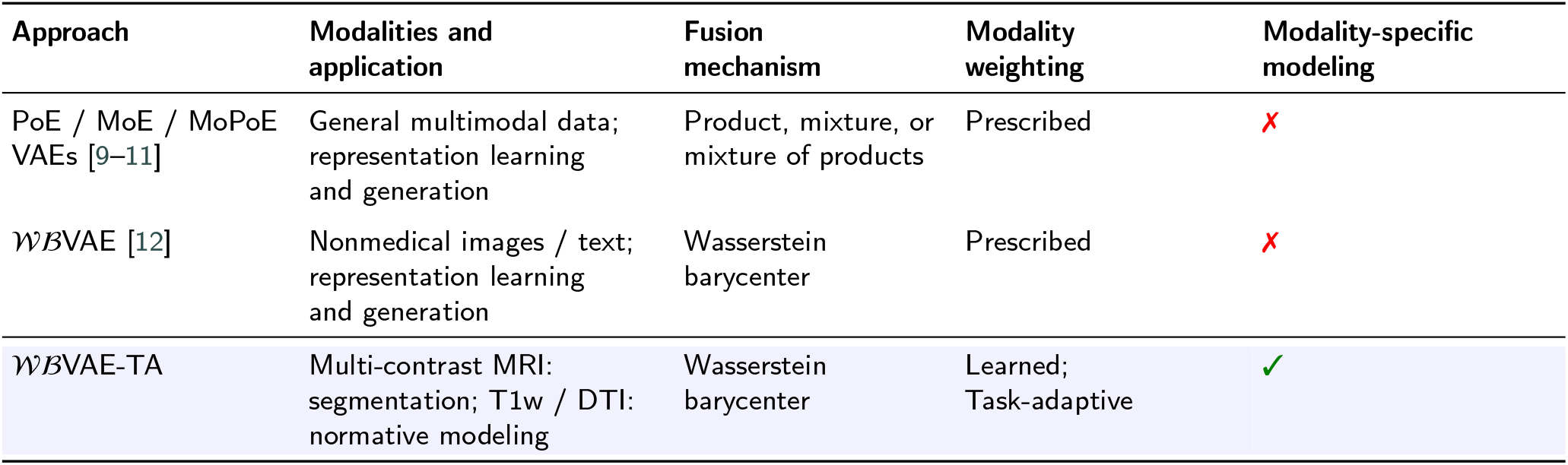
Representative multimodal methods compared by application and modeling design. This work proposes task-adaptive Wasserstein fusion with modality-specific conditioning for segmentation and normative modeling.

Motivated by these requirements, we develop task-adaptive Wasserstein barycentric fusion for multimodal medical imaging. Its first component, Adaptive *Wℬ* fusion, uses a Wasserstein barycenter with learnable global weights that determine each modality’s contribution. We denote the model with this fusion component alone as *Wℬ*VAE-A. All three variants use multilevel fusion. The second component adds learnable deterministic modality-specific embeddings to condition decoding features that propagate across network levels; these embeddings are exclusive to the full TA model. We refer to the resulting model as *Wℬ*VAE-TA, where TA denotes the full model, combining task-adaptive fusion with multilevel modality-specific decoding. Table 1 compares the proposed framework with prior multimodal VAE methods, highlighting its learned modality weighting and multilevel modality-specific conditioning.

We evaluate the proposed framework on two complementary medical applications: brain tumor segmentation with incomplete MRI inputs and normative modeling using structural and diffusion-derived measurements. Comparisons with established multimodal methods demonstrate improved performance across both tasks, while segmentation ablations quantify the consistent benefits of the proposed components. Specifically, learned weighting improves the mean Dice similarity coefficient (DSC) over *Wℬ*VAE by 2.72, 2.83, and 0.84 percentage points for enhancing tumor (ET), tumor core (TC), and whole tumor (WT), respectively. Adding the modality-specific conditioning component yields further improvements of 2.49, 0.95, and 0.42 percentage points over *Wℬ*VAE-A.

We broaden the barycentric fusion discussion in our prior *Wℬ*VAE [12] to general *αβ* divergences, whose parameters provide flexible control over probability mass allocation across modalities. Their barycenters generally lack closed-form solutions, even for Gaussian inputs, motivating Adaptive *Wℬ*fusion as a tractable approach to approximating selected mass-allocation behaviors. We learn the modality weights through the application objective while retaining closed-form diagonal Gaussian aggregation. To model modality-specific information alongside the fused shared representations, we further propose multiscale modality-specific decoding, using learnable deterministic modality embeddings to condition decoding features across network levels. We evaluate the resulting framework in brain tumor segmentation and normative modeling.

Our main contributions are:

1. **Adaptive** *Wℬ* **fusion (***Wℬ***VAE-A)**. We introduce task-learned modality weights within Wasserstein barycentric fusion, retaining closed-form diagonal Gaussian aggregation. Adaptive *Wℬ* can approximate selected mass-allocation behaviors of *αβ* barycenters.
2. **Multilevel task-adaptive fusion with modality-specific decoders**. We combine adaptive Wasserstein fusion at multiple network levels with modality-specific decoding pathways. Shared stochastic representations support modality-specific reconstruction, with learnable deterministic embeddings conditioning the initial decoder stage.
3. **Evaluation on two medical imaging tasks**. We assess *Wℬ*VAE-TA in brain tumor segmentation with missing modalities and multimodal normative modeling, with ablations of weighting and conditioning.

## 2. Related Work

### 2.1. Multimodal Representation Learning with VAEs

Multimodal VAEs can be roughly categorized into coordinated models [14–16] and joint models [9–13]. Coordinated models focus on aligning unimodal inference models and are not effective when some modalities are missing [17]. In contrast, joint models learn a joint distribution from all modalities, allowing for more robust handling of missing modalities by sampling from the joint latent space. This is particularly important in various medical domains, where certain modalities may be unavailable or infeasible to collect in clinical practice. Henceforth, we direct our main focus on this family of methods.

Joint models aggregate unimodal inference distributions to construct an approximate multimodal posterior. mVAE [9] uses a Product-of-Experts (PoE), while mmVAE [10] uses a Mixture-of-Experts (MoE). Their different concentration and coverage properties motivate alternatives such as MoPoE-VAE [11], which combines products over modality subsets using a mixture. The validity of a variational bound depends on the specified objective and variational distribution, rather than on the use of PoE alone. mmJSD [13] also considers shared and modality-specific information through a dynamic prior. Further variants explore this design space [18–20]. Our prior work [12] provided a barycentric interpretation of fusion; here we extend that framework to task-adaptive, multilevel shared representation learning with modality-specific conditioning.

### 2.2. Hierarchical Variational Autoencoders

Hierarchical VAEs, including Ladder VAE [21] and NVAE [22], increase generative-model capacity through multiple stochastic latent groups distributed across network levels. Ladder VAE recursively combines bottom-up evidence with top-down generative information, while NVAE uses a deep hierarchy of conditional latent distributions. These latent groups belong to the probabilistic generative model, have inferred posterior and prior distributions, and contribute corresponding KL terms to the variational objective.

Our model uses multiscale shared stochastic representations, with modality distributions fused at each level and the resulting samples supplied to modality-specific decoder pathways. Here, “multilevel” describes fusion and decoding across network resolutions, rather than a top-down hierarchy of conditional stochastic distributions as in Ladder VAE and NVAE. The modality embeddings are deterministic decoder parameters and do not themselves constitute additional stochastic latent groups.

### 2.3. Multimodal Learning in Medical Domains

The application of multimodal learning in medical image analysis focuses mainly on learning complementary cross-modal information and handling missing modalities. Following this avenue, [23] and [5] adapted the mVAE framework for multimodal brain MRI segmentation and multimodal normative modeling, respectively. [24] further adapted the MoPoE-VAE framework for multimodal normative modeling. Beyond generative models, discriminative models have also been explored for multimodal learning. In particular, mmFormer [25] employed a self-attention mechanism to capture cross-modal dependencies for the MRI segmentation task by attending to features extracted from the available modalities. It further encouraged consistency between outputs decoded from multimodal features and those decoded from unimodal features, improving robustness when some modalities are missing. Notably, this attention-based fusion can be viewed as a generalization of MoE [20], as it adaptively adjusts the contribution of each modality based on learned attention weights. Another line of work focuses on disentangling modality-invariant and modality-specific information to better exploit complementary modality cues. Approaches such as ShaSpec [26] and DC-Seg [27] explicitly separated shared anatomical structure from modality-specific appearance for the brain MRI segmentation task, enabling models to learn shared representations while preserving modality-specific information. Across all these approaches, randomly dropping modalities during training [28–30] has become a *de facto* standard for simulating missing-modality scenarios and encouraging models to remain robust when certain imaging sequences are unavailable. While effective in practice, modality dropout alone does not explicitly model the underlying fusion mechanism. The aforementioned methods are tailored to either segmentation or normative modeling. Our formulation applies to both generative and discriminative tasks, which we evaluate here.

## 3. Methodology

Without loss of generality, we consider a dataset 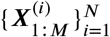 containing *N* independent and identically distributed (*i*.*i*.*d*.) samples, each of which consists of *M* modalities: 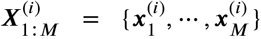. Assuming the multimodal data can be generated by some random process involving a joint latent variable *z*, the objective of a multimodal VAE is to maximize the evidence lower bound (*a*.*k*.*a*. ELBO) over all *M* modalities, under the *i*.*i*.*d*. assumption:

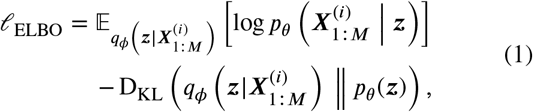

where 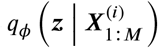is the fused approximate posterior and *p*_θ_(*z*) is the latent prior. Each modality has an encoding distribution 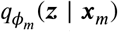and a decoder likelihood 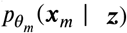. Assuming conditional independence given *z*, the joint likelihood factorizes as 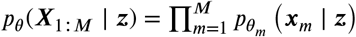. The true posterior is denoted *p*_θ_ (*z* ∣ *X*_1:*M*_) and is generally intractable. Sampling the prior supports unconditional generation; inference with incomplete observations instead requires an approximate posterior conditioned on the available modalities. We omit the subject index when it is not needed.

### 3.1. Multimodal VAE from a Barycentric View

Following the barycentric formulation [12], we construct the fused approximate posterior 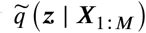from the unimodal encoding distributions:

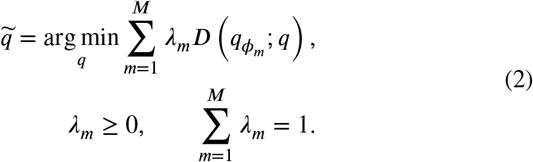

Here 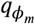 abbreviates 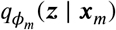, and *D*(⋅; ⋅) is a divergence or distance measure. The barycenter defines the variational distribution used in Eq. (1). It is parameterized and trained through the reconstruction and KL terms of that objective.

This barycentric formulation recovers a normalized weighted geometric pool and an arithmetic mixture by considering reverse and forward KL divergences, respectively [12]; the normalized weighted geometric pool differs from the standard unweighted PoE shown in panel (a), which sums component precisions. Exact derivations of the corresponding arithmetic and geometric pools are given in Appendix A.1. Notably, reverse and forward KL divergences exhibit distinct optimization behaviors: the reverse KL is mode-seeking (*i*.*e*., favoring bias), whereas the forward KL is mass-covering (*i*.*e*., favoring variance); See Fig. 1**a-b**. This information-theoretic property reveals the inherent trade-off between PoE and MoE and is consistent with empirical observations. Specifically, the performance of PoE tends to improve with an increasing number of modalities but may become biased toward certain dominant modalities. MoE retains a mixture of encoder distributions rather than necessarily producing a sharper single component.

Beyond the KL divergences discussed in [12], different choices of the distance measure *D*(⋅; ⋅) yield barycenters with distinct mass-allocation properties, providing a means to adjust the bias–variance trade-off (see Fig. 1**g**). This flexibility, however, must be balanced against computational tractability. For example, the *αβ* divergence is defined as

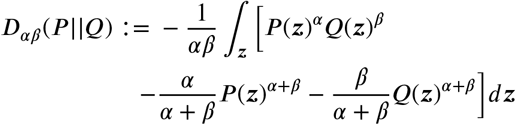

for *αβ*(*α* + *β*) ≠ 0. The KL cases are recovered : for normalized densities, (*α,β*) → (0 1) yields reverse KL, *D*_KL_ (*Q* ‖ *P*), whereas (*α,β*) → (1 0) yields forward KL, *D*_KL_ (*P* ‖ *Q*)),. Varying *α* and *β* changes how probability mass is allocated during multimodal fusion, allowing different bias–variance trade-offs (see Fig. 1**a-d**). However, generic *αβ* barycenters generally lack a closed-form solution, even when their input distributions are Gaussian. Numerical estimation remains possible, but introducing such an optimization into each fusion step complicates end-to-end training. These barycenters provide conceptual motivation for flexible mass allocation; their dedicated derivation and analysis are beyond the scope of this paper. The stationarity condition and special cases are provided in Appendix A.4.

Adaptive *Wℬ* broadens the flexibility of standard equal-weight Wasserstein fusion by allowing modality contributions to vary. These weights control the fused location and spread, providing a tractable means of approximating selected mass-allocation behaviors of *αβ* barycenters. The fused mean– standard-deviation pair lies in the convex hull of the component pairs. Consequently, fusion cannot produce coordinatewise standard deviations below the smallest component values or represent multimodal densities. This motivates the adaptive fusion formulation developed in § 3.2.

### 3.2. Adaptive Wasserstein Barycenter

We now derive the Wasserstein aggregation that implements this tractable control of fusion and then learn its modality weights through the task objective. Our closed-form implementation uses diagonal Gaussian encoders; the general covariance barycenter below is specified by an implicit equation. Without loss of generality, we first review the *p*-Wasserstein distance, which is the *p*-th root of the infimum of Kantorovich’s OT formulation [31]:

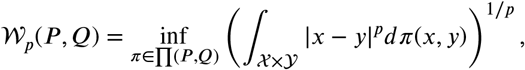

where the infimum is taken over the set of all transport plans *π* ∈ ∏(*P* , *Q*). In what follows, we consider the squared 2-Wasserstein distance 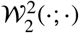instead of the widely used 1-Wasserstein distance (*a*.*k*.*a* earth mover distance), as 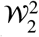 between two Gaussian distributions *N* (***µ***_1_, **Σ**_1_) and *N* (***µ***_2_, **Σ**_2_) can be solved analytically [32]:

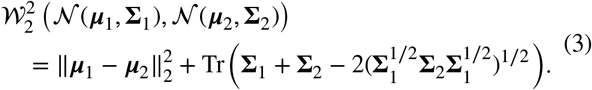

#### 3.2.1. Bures–Wasserstein Barycenter

The barycenter obtained by optimizing the squared 2-Wasserstein in Eq. (3) is the so-called Bures-Wasserstein barycenter [33]. Assuming unimodal encoding distributions 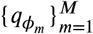 that are *d*-dimensional multivariate Gaussians 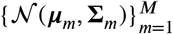denoting the mean and covariance of 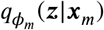, the resulting Bures-Wasserstein barycenter is also Gaussian-distributed 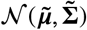:

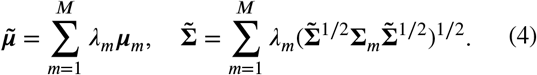

For general covariance matrices, Eq. (4) characterizes the barycenter implicitly [33]. For Gaussian posteriors with diagonal covariance, it simplifies to

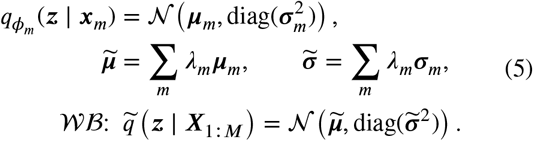

Here, *σ*_*m*_ ∈ ℝ^*d*^ contains standard deviations. The fused variances are therefore the elementwise squares of their weighted averages. Appendix A.2 contrasts this Gaussian Wasserstein solution with the Gaussian forms induced by the two KL orientations.

Equation (5) makes the additional flexibility explicit. Equal weights give the standard Wasserstein barycenter, whereas varying the weights shifts the fused mean and standard deviation toward different modalities while preserving closed-form aggregation. In Fig. 1**c-f**, suitable weights approximate changes in location and spread exhibited by numerical *αβ* barycenters. The remaining shape differences arise because the Gaussian Adaptive *Wℬ* family cannot represent arbitrary non-Gaussian target densities. The illustration therefore demonstrates selected mass-allocation behaviors accessible through adaptive weighting.

The illustration fits weights to numerical target densities to demonstrate this flexibility. In *Wℬ*VAE-A and *Wℬ*VAE-TA, we instead learn the weights through segmentation or generative losses, allowing modality contributions to adapt to the application objective without computing *αβ* barycenters during training.

#### 3.2.2. Adaptive Bures–Wasserstein Barycenter

To adapt multimodal fusion to medical imaging applications, we extend the original *Wℬ*VAE by learning global modality weights with the task objective. Segmentation and normative modeling place different demands on the shared representation. Learning the weights provides a means of optimizing the aggregation for each objective without changing the underlying Wasserstein geometry. In contrast to the target-fitted illustration, the training procedure does not compute *αβ* barycenters or optimize a discrepancy to them. At depth *l*, let 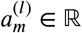be an unconstrained trainable logit, distinct from its normalized weight. Logits are learned separately at each depth and are not shared across depths. The weights are global across subjects:

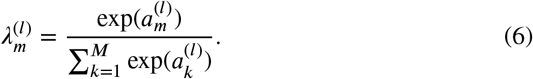

The model with learned modality weights is denoted

*Wℬ*VAE-A. The full model, *Wℬ*VAE-TA, also incorporates modality-specific conditioning across decoder levels. The relative utility of sequences such as T1ce and FLAIR motivates task-dependent weighting. The sensitivity of the fused Gaussian parameters to these logits is derived in Appendix A.3.

##### Fusion with an observed modality subset

The natural extension to a nonempty observed set *S* ⊆ {1, … , *M*} restricts the fusion objective to that set:

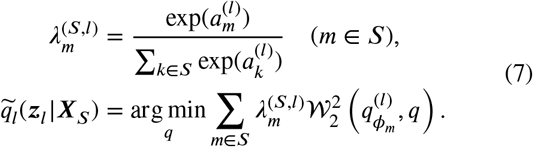

Here 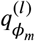denotes the modality-*m* encoding distribution at depth. This specifies a normalized subset fusion operator: a singleton set recovers its unimodal encoder distribution, and the empty set is excluded. During segmentation training, one modality subset is sampled uniformly from all 15 nonempty subsets of the four MRI modalities at each iteration. In the following remark, we consider one fusion depth and suppress its index *l* for readability.

##### Remark (Task-directed weight gradients)

Fix the encoders and decoders. For an observed set *S*, let *v*_*m*_ = (***µ***_*m*_, *σ*_*m*_) and 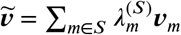. For a differentiable task loss *L* to be minimized that depends on the logits only through fusion,

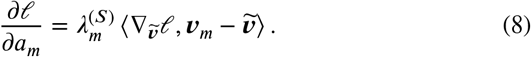

The identity follows from the softmax derivative and the chain rule. It relates the task gradient to the displacement of each modality’s mean–standard-deviation pair from the fused pair, describing how the task objective guides the learned weights while retaining closed-form fusion. A derivation and its expected-loss interpretation are given in Appendix A.3.1.

### 3.3. Modality-Conditioned Multilevel Decoding

To provide modality-dependent modeling capacity alongside shared structure, we augment shared stochastic representations with learnable modality-specific embeddings 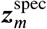 in Fig. 2. From a generative perspective, shared fusion a^*m*^nd modality-specific conditioning play complementary roles. Wasserstein barycentric aggregation constructs shared latent distributions from the available modalities, whereas the embeddings condition the observation models that decode these representations. For the shared latent collection 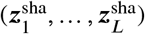, modality *m* is modeled through 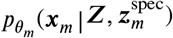. The decoder parameters θ_*m*_ define the modality-specific observation models, while the embedding supplies an explicit conditioning signal within each multilevel decoding pathway. Because the embeddings are deterministic model parameters shared across subjects, rather than subject-specific random variables with inferred posterior distributions, they do not introduce additional latent probability distributions.

**Figure 2.**
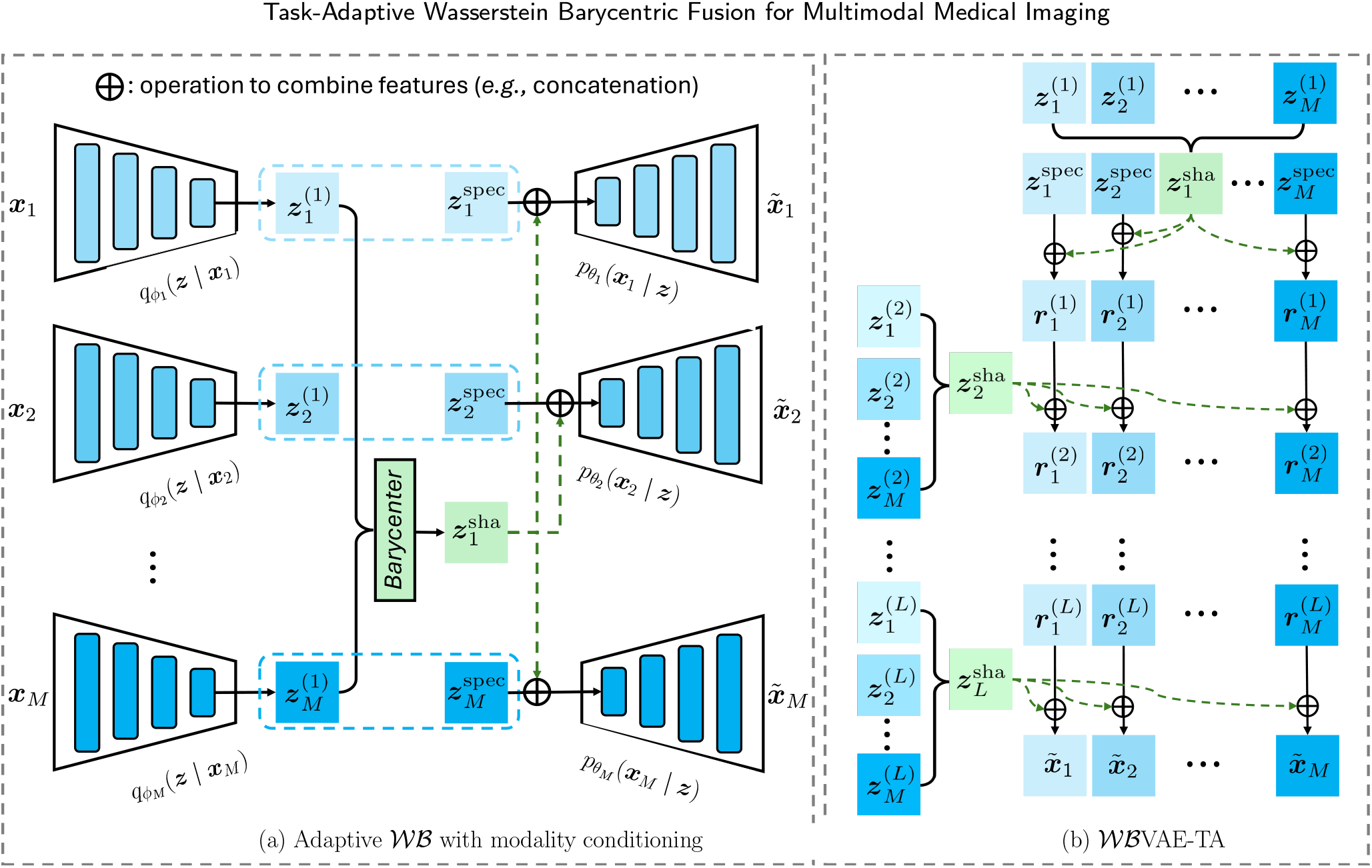
The overview of the proposed method for multimodal representation learning: **(a)** Adaptive *Wℬ* fusion with modality-specific embeddings and **(b)** the complete multilevel *Wℬ*VAE-TA design. Panel (a) illustrates the fusion and conditioning components rather than the weighting-only ablation. The notation 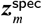 denotes the learnable modality-specific embedding for modality *m*. Concatenated features pass through decoder blocks; the next stage produces 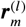 from 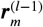and the shared latent at stage *l*. The stage indexing is specified in Eq. (9). At stage *l*, 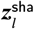and 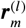denote the fused shared latent vectors and the modality-*m* feature vector that combines modality-invariant and modality-specific components, respectively.

At each encoder stage *l*, Wasserstein fusion acts on the unimodal encoding *distributions*, producing 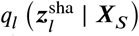A shared latent sample is then drawn from this fused distribution. Let 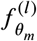denote a deterministic decoder block and [⋅, ⋅] feature concatenation. The multilevel decoder is written as

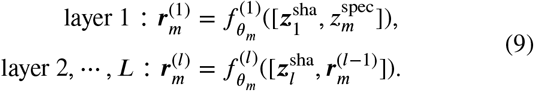

The output head maps the final features to the reconstructed modalities 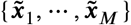. The modality-specific embedding enters at the first decoder stage and is propagated through the modality-conditioned feature state 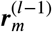. At each sub-sequent stage, this state is combined with^*m*^ the shared latent representation at the corresponding resolution. The resulting recurrence enables modality-dependent interpretation of shared information across decoder levels without independently injecting the embedding at every stage. Because the stated observation models use modality-specific decoder parameters, a constant embedding concatenated before an affine block can be absorbed into that block’s bias when its contribution is spatially constant. Any additional expressive role depends on the actual decoder operations and parameter sharing.

Let *q*_*ϕ*_(*Z* ∣ *X*_*S*_) denote the joint variational distribution over the shared latent collection. For a sampled observed subset *S*, inference uses only *X*_*S*_, while the reconstruction term includes all four MRI modalities, including those omitted from the input. The corresponding variational objective is

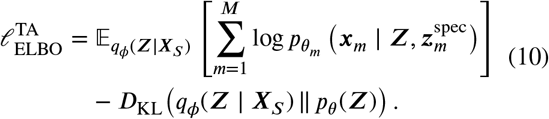

No separate KL term for 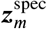arises because it is a deterministic parameter. Instead^*m*^, each embedding is optimized through the expected reconstruction term, while the KL term regularizes the shared stochastic representations. In our implementation, the posterior factorizes across latent levels as 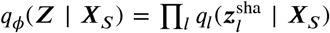, and the prior is 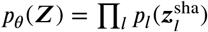 with *p*_*l*_ = *N*(**0, *I***) at every level. The joint KL therefore reduces to a sum of per-level KL terms. We use *L* for the application loss to be minimized and 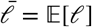for its expected training objective, consistent with the weight-gradient identity in Eq. (8). For normative modeling, 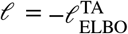; segmentation additionally includes Dice and cross-entropy losses. The application-specific network architectures are provided in Appendix B.

## 4. Experiments

To validate the effectiveness of the proposed method, we conducted experiments on two tasks that commonly involve multiple imaging modalities: *i)* multimodal brain tumor segmentation (§ 4.1) and *ii)* multimodal normative modeling (§ 4.2). Given the fundamental differences between these tasks, their experimental designs vary slightly; we therefore present them separately.

### 4.1. Multimodal Brain Tumor Segmentation

#### 4.1.1. Experimental Setup

Following previous literature [23, 25, 26], we validated our method on Brain Tumor Segmentation (BraTS) Challenge 2018 dataset [1], which consists of 285 multimodal MRI scans with four modalities (*M* = 4): T1w, T1ce, T2w, and FLAIR. The dataset provides voxel-wise annotations for tumor subregions, including enhancing tumor (ET), tumor core (TC), and edema (ED). Following standard practice, we report Dice Similarity Coefficient (DSC) on three nested evaluation regions: whole tumor (WT), tumor core (TC), and enhancing tumor (ET). The dataset split follows mmFormer [25], with 199 subjects for training, 29 for validation, and 57 for testing, for a fair comparison. Following mmFormer [25] and ShaSpec [26], the original 240 × 240 × 155 MRI volumes were cropped into 128 × 128 × 128 patches for training.

To support the segmentation task, an additional segmentation decoder is added to *Wℬ*VAE-TA that shares the same architecture as the other VAE decoders. Its output represents tumor labels rather than modality reconstruction, while the reconstruction decoders parameterize the image likelihoods (see Table B.1). The segmentation decoder is trained jointly with the other probabilistic decoders by minimizing *ℓ*, which combines Dice and cross-entropy losses with the negative multimodal ELBO in Eq. (10), using weights of 0.5, 0.5, and 1.0, respectively. We trained the model for 1000 epochs using AdamW [34] with an initial learning rate of 3 × 10^−4^, a cosine learning-rate schedule, and a batch size of 1 on an NVIDIA Tesla A100 GPU with 40 GB of memory. To mitigate overfitting, we applied data augmentation during training, including random flipping, cropping, and intensity shifts.

##### Baselines

We compare against U-HVED [23], mmFormer [25], ShaSpec [26], and DC-Seg [27], selected to represent complementary approaches to multimodal brain tumor segmentation with missing modalities. U-HVED provides a probabilistic multimodal VAE baseline with PoE fusion, allowing comparison with the proposed Wasserstein barycentric aggregation. mmFormer provides an attention-based alternative for learning interactions among available modalities. ShaSpec and DC-Seg explicitly model shared and modality-specific information, making them relevant comparisons for our modality-specific conditioning design. Together, these baselines assess the proposed framework against probabilistic fusion, attention-based fusion, and shared/specific representation learning under complete and incomplete inputs. mmFormer, ShaSpec, and DC-Seg use their official codebases. U-HVED is re-implemented using the same encoder–decoder architecture and configuration as *Wℬ*VAE, *Wℬ*VAE-A, and *Wℬ*VAE-TA, as described in Appendix B, while retaining its original training objective. We compare the model complexity between the proposed method and baselines in Appendix C.

#### 4.1.2. Performance on Brain Tumor Segmentation

Table 2 reports segmentation performance under complete and incomplete modality settings across all 15 combinations of the four MRI modalities. Compared with the attention-based mmFormer, the proposed method improved the average DSC by 2.31, 2.73, and 0.76 on the segmentation of ET, TC, and WT, respectively, across all possible combinations of modalities. Similarly, our method improved the average DSC over PoE-based U-HVED by 8.38 (ET), 7.04 (TC), and 3.70 (WT) across all possible combinations of modalities. Our method significantly outperforms the compared baselines in the comparisons marked by the Wilcoxon tests in the table. Fig. 3 describes sensitivity to modality availability; its standard deviations are not confidence intervals. Compared with ShaSpec and DC-Seg, the reported average improvements are 2.16 and 2.87 points for ET, 2.14 and 1.38 for TC, and 0.80 and 0.59 for WT, respectively. Individual modality configurations do not uniformly favor the proposed method.

**Table 2.** Performance on multimodal segmentation, measured by Dice similarity coefficient (DSC) [%] for all modality combinations. ET, TC, and WT denote enhancing tumor, tumor core, and whole tumor, respectively. • and ∘ denote available and missing modalities. The highest average DSC for each tumor region is highlighted in **bold**.

| Method |  | 1 modality |  |  |  | 2 modalities |  |  |  |  |  | 3 modalities |  |  |  | 4 modalities | Average |
| --- | --- | --- | --- | --- | --- | --- | --- | --- | --- | --- | --- | --- | --- | --- | --- | --- | --- |
|  | FLAIR | ● | ○ | ○ | ○ | ● | ● | ● | ○ | ○ | ○ | ● | ● | ● | ○ | ● |  |
|  | T1ce | ○ | ● | ○ | ○ | ● | ○ | ○ | ● | ● | ○ | ● | ● | ○ | ● | ● |  |
|  | T1w | ○ | ○ | ● | ○ | ○ | ● | ○ | ● | ○ | ○ | ● | ○ | ● | ● | ● |  |
|  | T2w | ○ | ○ | ○ | ● | ○ | ○ | ● | ○ | ● | ● | ○ | ● | ● | ● | ● |  |
| ET | U-HVED [23] (MICCAI'19) | 28.40 | 66.31 | 25.58 | 32.45 | 70.27 | 38.34 | 37.72 | 71.96 | 68.46 | 39.26 | 71.41 | 70.40 | 41.50 | 74.41 | 72.55 | 53.93 |
|  | mmFormer [25] (MICCAI'22) | 41.37 | 73.72 | 35.26 | 39.84 | 75.15 | 44.41 | 44.87 | 72.83 | 75.05 | 45.05 | 76.42 | 76.87 | 48.69 | 74.49 | 76.00 | 60.00 |
|  | ShaSpec [26] (CVPR'23) | 38.69 | 75.61 | 35.34 | 40.91 | 76.09 | 42.68 | 48.87 | 75.99 | 74.83 | 45.29 | 77.66 | 73.60 | 45.13 | 76.35 | 75.15 | 60.15 |
|  | DC-Seg [27] (MICCAI'25) | 39.97 | 72.78 | 38.77 | 41.90 | 73.04 | 46.55 | 44.99 | 75.87 | 73.32 | 45.58 | 72.58 | 72.87 | 48.28 | 72.75 | 72.42 | 59.44 |
|  | WBVAE-TA | 37.05 | 76.69 | 44.58 | 42.44 | 77.75 | 46.37 | 46.36 | 77.67 | 78.02 | 49.45 | 77.48 | 77.84 | 51.03 | 76.09 | 75.76 | 62.31* |
| TC | U-HVED [23] (MICCAI'19) | 58.60 | 74.80 | 57.82 | 62.00 | 79.28 | 67.76 | 67.22 | 78.50 | 77.99 | 66.22 | 80.01 | 79.24 | 68.59 | 79.12 | 79.74 | 71.79 |
|  | mmFormer [25] (MICCAI'22) | 67.96 | 80.48 | 68.01 | 65.80 | 82.60 | 71.01 | 68.98 | 82.45 | 82.51 | 68.95 | 83.18 | 82.41 | 71.13 | 83.22 | 82.83 | 76.10 |
|  | ShaSpec [26] (CVPR'23) | 68.29 | 81.94 | 65.65 | 67.54 | 83.76 | 71.78 | 70.82 | 82.79 | 83.22 | 69.24 | 83.76 | 83.94 | 70.76 | 83.30 | 83.61 | 76.69 |
|  | DC-Seg [27] (MICCAI'25) | 68.38 | 81.59 | 69.76 | 67.78 | 83.75 | 73.35 | 70.93 | 83.34 | 83.82 | 70.42 | 84.05 | 83.61 | 72.70 | 84.34 | 84.00 | 77.45 |
|  | WBVAE-TA | 69.73 | 82.09 | 70.73 | 71.04 | 82.77 | 74.36 | 75.37 | 83.93 | 84.46 | 74.04 | 83.99 | 84.64 | 75.78 | 84.76 | 84.82 | 78.83* |
| WT | U-HVED [23] (MICCAI'19) | 83.30 | 70.79 | 69.86 | 80.10 | 87.90 | 87.34 | 88.18 | 76.93 | 83.74 | 84.17 | 88.65 | 89.21 | 89.08 | 85.18 | 89.55 | 83.60 |
|  | mmFormer [25] (MICCAI'22) | 88.44 | 76.92 | 77.09 | 84.55 | 89.47 | 89.55 | 89.80 | 80.91 | 86.84 | 86.83 | 89.77 | 90.17 | 90.13 | 87.30 | 90.31 | 86.54 |
|  | ShaSpec [26] (CVPR'23) | 88.05 | 77.12 | 78.04 | 84.51 | 89.70 | 89.32 | 89.88 | 80.65 | 86.21 | 86.37 | 90.06 | 90.29 | 90.08 | 86.71 | 90.54 | 86.50 |
|  | DC-Seg [27] (MICCAI'25) | 87.70 | 78.28 | 79.47 | 83.50 | 89.86 | 89.81 | 89.58 | 81.68 | 86.33 | 86.46 | 90.19 | 90.23 | 90.11 | 87.10 | 90.42 | 86.71 |
|  | WBVAE-TA | 89.40 | 79.66 | 79.48 | 85.49 | 89.84 | 90.20 | 90.19 | 81.83 | 86.92 | 87.38 | 90.73 | 90.09 | 90.22 | 87.40 | 90.67 | 87.30* |

\* $p < 0.05$ ; with Wilcoxon signed rank test to the second-best models.

**Figure 3.**
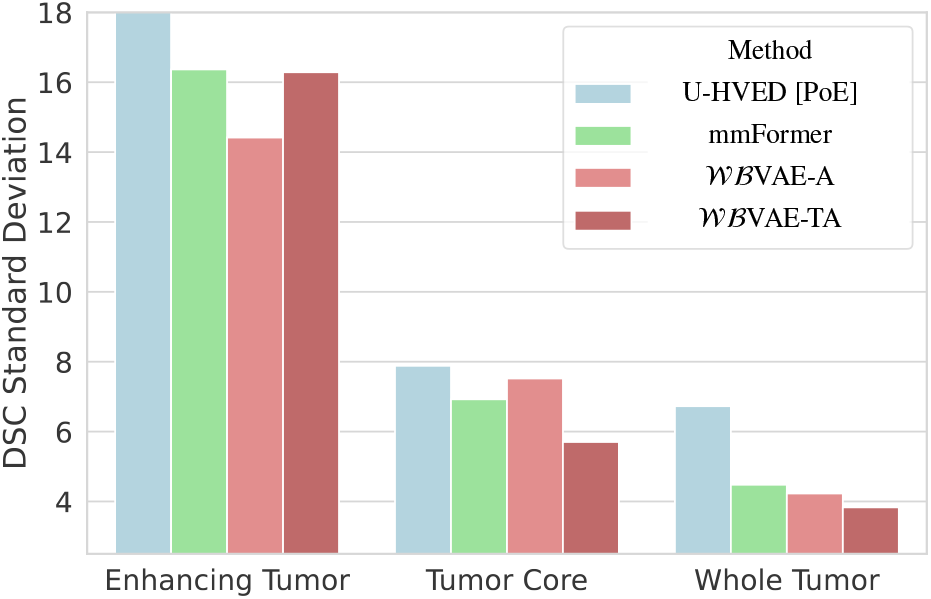
Standard deviation of DSC across the 15 modality combinations. This measures variation across input configurations, not uncertainty across subjects or independent training seeds. Lower variation alone does not establish improved posterior probability-mass coverage.

#### 4.1.3 Qualitative Analysis

In Fig. 4, we present predictions for one subject under four modality-availability scenarios. With T1w alone, *Wℬ*VAE-TA achieves WT/TC/ET DSC of 0.913/0.842/0.638. Its WT score is favorable, but ShaSpec achieves a higher TC score (0.894) and DC-Seg a higher ET score (0.688). With all four inputs, the proposed model achieves 0.953/0.969/0.891. This illustrative case does not establish consistent superiority across subjects or explain the latent mechanism underlying the differences.

**Figure 4.**
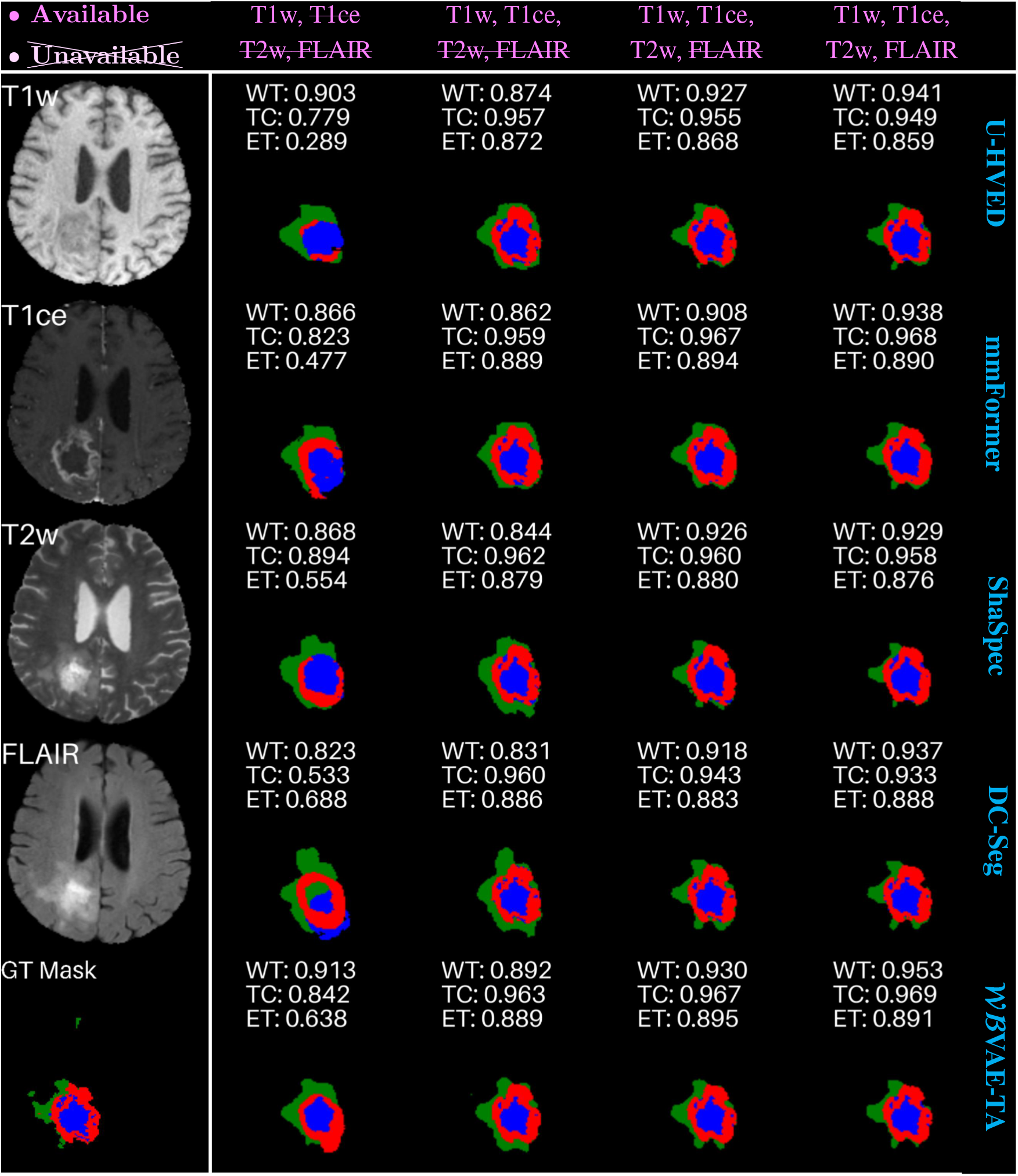
Qualitative comparison of brain tumor segmentation results obtained from different methods (rows) across different multimodal input configurations: {T1w}, {T1w, T1ce}, {T1w, T1ce, T2w}, {T1w, T1ce, T2w, FLAIR}. Sub-tissue classes are color-coded: 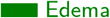, 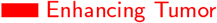, 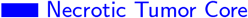.

#### 4.1.4. Ablation Study

To assess the contribution of each component, we performed ablation studies on different model variants (*i.e*., *Wℬ*VAE, *Wℬ*VAE-A, and *Wℬ*VAE-TA). All three variants use multilevel fusion. *Wℬ*VAE uses equal fusion weights,

whereas *Wℬ*VAE-A learns the fusion weights. Learnable deterministic modality-specific embeddings are included only in *Wℬ*VAE-TA. The results are shown in Table 3. The UHVED results in Table 2 provide a complementary controlled comparison with the WB-based variants using the same multi-level encoder–decoder architecture and configuration. Specifically, averaged across all modality combinations, *Wℬ*VAE-A consistently improved performance over *Wℬ*VAE, increasing average DSC from 57.10 to 59.82 on ET (+2.72), from 75.05 to 77.88 on TC (+2.83), and from 86.04 to 86.88 on WT (+0.84). These gains are consistent with a benefit from learned Wasserstein fusion under the evaluated modality configurations. Extending *Wℬ*VAE-A with modality-specific embedding conditioning yields further average DSC improvements of 2.49, 0.95, and 0.42 percentage points for ET, TC, and WT, respectively. The benefits of modality-specific embedding conditioning are more pronounced in challenging settings with severe modality missingness. For example, with only T1ce available, the DSC of ET improved from 69.99 to 76.69, and with only T1w available, the DSC of TC improved from 68.40 to 70.73 (see Table 3). For two-modality inputs, *Wℬ*VAE-TA continued to improve the DSC of ET, *e.g*., the DSC of ET improved from 46.86 to 49.45 with T1w and T2w modalities; whereas in more complete settings (three or four modalities), gains tend to be smaller and occasionally fluctuate. The learned modality weights, summarized across network levels, are reported in Appendix D, where T1ce and FLAIR receive higher weights than other modalities, consistent with their relevance to delineating enhancing tumor and peritumoral edema, respectively.

**Table 3.** Ablations on the multimodal segmentation task. Dice similarity coefficient (DSC) [%] is employed for evaluation with every combination of modalities. • and ∘ denote available and missing modalities, respectively. The underscript + denotes the performance gain when moving from one model to the next (left to right).

| Modalities |  |  |  |  | Enhancing Tumor (ET) |  |  | Tumor Core (TC) |  |  | Whole Tumor (WT) |  |  |
| --- | --- | --- | --- | --- | --- | --- | --- | --- | --- | --- | --- | --- | --- |
| FLAIR | T1ce | T1w | T2w |  | WBVAE | WBVAE-A | WBVAE-TA | WBVAE | WBVAE-A | WBVAE-TA | WBVAE | WBVAE-A | WBVAE-TA |
| ● | ○ | ○ | ○ |  | 39.89 | 41.98 | 37.05 | 66.55 | 66.92 | 69.73 | 87.92 | 88.53 | 89.40 |
| ○ | ● | ○ | ○ |  | 67.31 | 69.99 | 76.69 | 80.36 | 82.35 | 82.09 | 75.99 | 77.35 | 79.66 |
| ○ | ○ | ● | ○ |  | 36.26 | 36.80 | 44.58 | 60.76 | 68.40 | 70.73 | 76.79 | 79.14 | 79.48 |
| ○ | ○ | ○ | ● |  | 41.23 | 43.11 | 42.44 | 67.29 | 68.16 | 71.04 | 84.07 | 85.36 | 85.49 |
| ● | ● | ○ | ○ |  | 72.20 | 75.96 | 77.75 | 82.47 | 85.04 | 82.77 | 89.58 | 90.31 | 89.84 |
| ● | ○ | ● | ○ |  | 44.54 | 46.51 | 46.37 | 69.35 | 71.07 | 74.36 | 89.10 | 89.78 | 90.20 |
| ● | ○ | ○ | ● |  | 45.84 | 48.09 | 46.36 | 69.05 | 71.71 | 75.37 | 89.67 | 90.30 | 90.19 |
| ○ | ● | ● | ○ |  | 66.92 | 73.51 | 77.67 | 81.57 | 84.34 | 83.93 | 79.76 | 80.88 | 81.83 |
| ○ | ● | ○ | ● |  | 70.68 | 69.95 | 78.02 | 82.07 | 84.84 | 84.46 | 85.69 | 86.76 | 86.92 |
| ○ | ○ | ● | ● |  | 43.38 | 46.86 | 49.45 | 67.51 | 71.52 | 74.04 | 85.82 | 86.95 | 87.38 |
| ● | ● | ● | ○ |  | 70.16 | 75.75 | 77.48 | 82.48 | 85.26 | 83.99 | 89.85 | 90.34 | 90.73 |
| ● | ● | ○ | ● |  | 69.71 | 75.46 | 77.84 | 82.85 | 85.41 | 84.64 | 90.08 | 90.09 | 90.09 |
| ● | ○ | ● | ● |  | 45.23 | 49.53 | 51.03 | 68.90 | 72.30 | 75.78 | 89.82 | 90.38 | 90.22 |
| ○ | ● | ● | ● |  | 70.84 | 70.26 | 76.09 | 82.12 | 85.28 | 84.76 | 86.18 | 86.93 | 87.40 |
| ● | ● | ● | ● |  | 72.36 | 73.55 | 75.76 | 82.47 | 85.64 | 84.82 | 90.26 | 90.04 | 90.67 |
| Average |  |  |  |  | 57.10 | 59.82<sub>+2.72</sub> | 62.31<sub>+2.49</sub> | 75.05 | 77.88<sub>+2.83</sub> | 78.83<sub>+0.95</sub> | 86.04 | 86.88<sub>+0.84</sub> | 87.30<sub>+0.42</sub> |

### 4.2. Multimodal Normative Modeling

#### 4.2.1. Experimental Setup

Following [5, 24], we validated our method using UK Biobank [35] and Alzheimer’s Disease Neuroimaging Initiative (ADNI) [36] datasets. For both datasets, we used two modalities (*M* = 2): *i)* T1w MRI, from which grey-matter volumes were extracted for 66 cortical (Desikan-Killiany atlas) and 16 subcortical brain regions, and *ii)* Diffusion Tensor Imaging (DTI), yielding Fractional Anisotropy (FA) and Mean Diffusivity (MD) measurements for 35 white-matter tracts (Johns Hopkins University atlas). We consider both modalities in the following experiments because they provide complementary information about individual variation, allowing normative modeling to characterize deviations that may be less apparent when either modality is considered alone. This choice also builds on prior multimodal work and facilitates a direct comparison with existing findings [5, 24]. Unlike segmentation, which uses an additional segmentation decoder, normative modeling evaluates the generative variants without a segmentation head.

*Baselines*. We compare against mVAE [9], weighted-mVAE [5], mmVAE [10], mmJSD [13], and MoPoE-VAE [24], selected to cover complementary multimodal generative formulations and established approaches to normative modeling. mVAE and mmVAE provide PoE- and MoE-based reference models, respectively, for comparing Wasserstein barycentric fusion with product and mixture aggregation. Weighted-mVAE provides a modality-weighted PoE comparison previously applied to normative modeling, making it relevant to our learned weighting mechanism. mmJSD provides a comparison based on a multimodal Jensen–Shannon objective and a dynamic prior, while MoPoE-VAE combines products over modality subsets within a mixture and connects our evaluation to prior multimodal normative modeling work. Together, these baselines assess whether the proposed formulation improves data modeling and deviation-based evaluation across different fusion strategies. We additionally report *Wℬ*VAE-A as a within-framework comparison to *Wℬ*VAE-TA.

Given the limited size of the ADNI cohort, we adopted a two-stage transfer-learning strategy to reduce data requirements while accommodating differences in population and acquisition characteristics. We first pretrained the model on the larger UK Biobank cohort to learn shared representations, then fine-tuned it on ADNI. For pretraining, the UK Biobank development cohort comprised 4,797 healthy participants, including 564 reserved for validation. The model was trained for 1,000 epochs using the Adam optimizer, a batch size of 256, and an initial learning rate of 10^−5^. We evaluated the pretrained model on a separate held-out test set of 847 healthy UK Biobank participants. We then fine-tuned the model for 100 epochs using 93 healthy ADNI participants. Given the small sample size, we used three-fold cross-validation and reported performance averaged across folds. Disease-detection performance was subsequently evaluated on 170 ADNI participants with mild cognitive impairment (MCI; *n* = 138) or Alzheimer’s disease (AD; *n* = 32), classified according to Clinical Dementia Rating (CDR) criteria.

Most prior work [5, 24] evaluated normative models using a single metric, namely the significance ratio [5], defined as the ratio between the true positive rate (TPR) and the false positive rate (FPR). To determine the true positives or false positives, one computes the Mahalanobis distance between each subject’s latent representation and the reference healthy distribution. Distances are evaluated for subjects from both the disease cohort and a held-out healthy cohort, with the reference healthy cohort typically corresponding to the training cohort [5, 24]. We can then calculate the TPR and FPR by counting individuals whose deviations were significantly different from the healthy training cohort, determined by *p* < 0.001. However, we argue that relying solely on the significance ratio can be limiting. This limitation arises from three factors: *i)* the significance ratio is unbounded (*i.e*., it has no fixed lower or upper bound), which complicates interpretation and comparison across settings; *ii)* it becomes highly unstable when the FPR is small, such that minor fluctuations in FPR can induce large swings in the ratio, particularly when the held-out healthy cohort is small; and *iii)* it provides no direct assessment of whether the VAE has learned a faithful representation of the underlying multimodal data distribution.

To address these issues, we complemented the significance ratio with three additional metrics to enable a more comprehensive evaluation. First, we included **precision**, the proportion of flagged subjects who belong to the disease cohort. This value depends on the disease prevalence in the evaluation cohort and should not be interpreted as a population-level clinical predictive value. Second, we used **balanced accuracy**, defined as the mean of sensitivity and specificity, to jointly capture detection performance on both diseased and healthy cohorts and to provide a bounded, interpretable measure. Finally, to assess representation fidelity beyond detection outcomes, we estimated the data **log-likelihood** log *p*(*X*_1:*M*_) under the learned multimodal VAE, indicating how well the model approximated the multimodal data distribution and whether its latent representations were consistent with the observed data. Following [11, 13], the reported evaluation uses *K* = 10 importance samples. The normalized estimator of the log-likelihood is

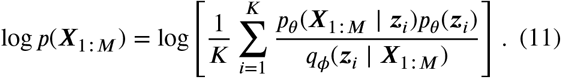

After healthy-cohort fitting, disease detection uses each subject’s encoding latent representation to compute its Mahalanobis deviation from the reference healthy distribution, followed by the reported *p* < 0.001 decision rule. This is a deviation-based evaluation, not a separately trained disease-classification head.

#### 4.2.2. Performance on Normative Modeling

Table 4 reports estimated data log-likelihood on UK Biobank and ADNI, together with deviation-based disease-detection metrics on ADNI. Notably, our *Wℬ* VAE-TA out-performed all baseline methods by a large margin in estimated data log-likelihood on both UK Biobank and ADNI, indicating a strong capacity to model the multimodal data distribution. The proposed *Wℬ* VAE-TA also outperformed the design variant *Wℬ* VAE-A in all the metrics. The improvement in estimated data log-likelihood supports the combined multilevel conditioning design, which complements the shared representations with an explicit modality-dependent decoding pathway. In the downstream disease detection task on ADNI, *Wℬ* VAE-TA achieved the highest reported significance ratio, precision, and balanced accuracy among the evaluated models. These experiments use complete inputs and do not establish missing-modality robustness for normative modeling.

**Table 4.** Performance on normative modeling with both T1w and DTI available. We report the estimated data log-likelihood log *p*(*X*_1:*M*_) on UK Biobank and significance ratio (sig. ratio), precision, balanced accuracy (bal. accuracy), and log *p*(*X*_1:*M*_) on ADNI.

| Model | UK Biobank<br>$\log p(\mathbf{X}_{1:M}) \uparrow$ | ADNI | | | |
| --- | --- | --- | --- | --- | --- |
| | | sig. ratio ↑ | precision ↑ | bal. accuracy ↑ | $\log p(\mathbf{X}_{1:M}) \uparrow$ |
| mVAE [9] | -4.222 | 1.464 | 0.863 | 0.589 | -5.052 |
| weighted-mVAE [5] | -4.191 | 1.200 | 0.885 | 0.535 | -4.932 |
| mmVAE [10] | -4.648 | 1.449 | 0.887 | 0.585 | -5.588 |
| mmJSD [13] | -5.220 | 1.438 | 0.882 | 0.566 | -6.264 |
| MoPoE-VAE [24] | -4.219 | 1.491 | 0.894 | 0.586 | -5.671 |
| $\mathcal{WBVAE}$-A | -4.180 | 1.613 | 0.895 | 0.591 | -4.989 |
| $\mathcal{WBVAE}$-TA | **-3.146** | **1.803** | **0.907** | **0.599** | **-3.821** |

#### 4.2.3. Separation of Disease Stages

We assessed the model’s ability to distinguish disease stages (CU, MCI, and AD) by comparing latent deviation scores quantified as Mahalanobis distances from the healthy cohort distribution (see Fig. 5). Overall, all methods showed the expected monotonic increase in deviation scores across disease stages (CU ≺ MCI ≺ AD). However, the degree of separation and statistical consistency varies: most methods yield overlapping distributions between CU and MCI (often marked as non-significant), indicating limited sensitivity to early-stage deviation. In contrast, *Wℬ* VAE-TA exhibited the clearest stage-wise stratification, with a well-ordered shift in medians from CU to MCI to AD and comparatively tighter dispersion. *Wℬ* VAE-TA also improved separability between CU and AD and between adjacent stages (CU vs. MCI and MCI vs. AD), with nominally significant CU–MCI separation. These stage comparisons suggest differences in latent deviations across clinical groups; further validation is needed to establish clinical diagnostic utility.

**Figure 5.**
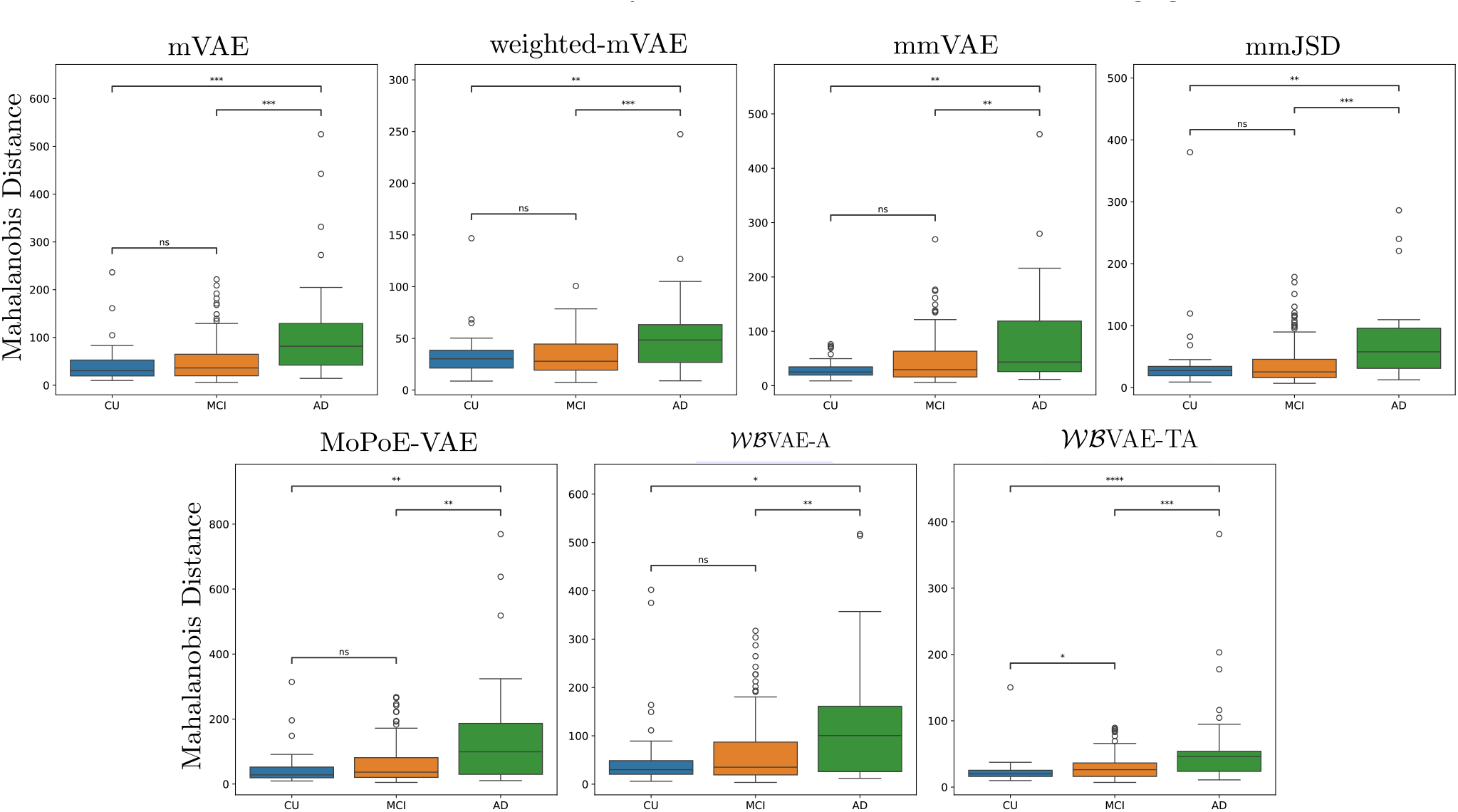
Comparison of the latent deviation scores across different clinical stages (CU, MCI, and AD). Statistical annotations were generated using Welch’s t-tests between pairs of CU, MCI, and AD groups. ns: 0.05 < *p* ≤ 1, ∗: 0.01 < *p* ≤ 0.05, ∗∗: 0.001 < *p* ≤ 0.01, ∗∗∗: 0.0001 < *p* ≤ 0.001, ∗∗∗∗: *p* ≤ 0.0001.

## 5. Discussion

The broader barycentric view connects the flexibility of multimodal fusion to the allocation of probability mass across modality distributions. Although *αβ* divergences offer flexible control over this allocation, their barycenters generally lack closed-form solutions. Adaptive *Wℬ* provides a tractable way to broaden equal-weight Wasserstein fusion by adjusting modality contributions. The Gaussian formulation makes these adjustments explicit through the fused location and spread, and Fig. 1 illustrates selected behaviors that approximate numerical *αβ* barycenters. The illustration uses target-fitted weights, whereas our models learn weights directly through the application objective. The weight-gradient identity in Eq. (8) describes how the task loss guides these updates through each modality’s displacement from the fused mean–standard-deviation pair.

Adaptive fusion and modality-specific decoding address complementary modeling needs. In *Wℬ* VAE-A, learned weights determine the contribution of each modality to the shared stochastic representation. In *Wℬ* VAE-TA, multilevel fusion supplies shared stochastic representations to modality-specific decoding pathways, with learnable embeddings conditioning the initial decoder stage. This design provides a common framework for segmentation and normative modeling, with the fusion weights and decoding parameters optimized through their respective objectives.

The segmentation ablations show consistent benefits from learned weighting and modality-specific embedding conditioning. Across the 15 modality combinations, *Wℬ* VAE-A improves average DSC over equal-weight *Wℬ* VAE. Extending *Wℬ* VAE-A with modality-specific embedding conditioning yields further average DSC improvements of 2.49, 0.95, and 0.42 percentage points for ET, TC, and WT, respectively. The learned weights favor T1ce and FLAIR, consistent with the complementary information these sequences provide for enhancing tumor and peritumoral edema. Using the same encoder–decoder architecture and configuration provides a controlled comparison of U-HVED with the WB-based variants under a shared backbone. The results demonstrate the advantage of our WB-based models over the PoE-based U-HVED baseline. Together, these results support adapting modality contributions and incorporating modality-specific information when learning from incomplete MRI inputs.

Normative modeling examines the same framework under a generative objective. The improved estimated data log-likelihood and ADNI deviation-based detection scores support its utility for modeling healthy variation and identifying departures from that reference. The nearly balanced T1w and DTI weights differ from the segmentation weighting pattern, illustrating how modality contributions depend on the application and input representations. These experiments use complete inputs, and the ADNI cohort is small, motivating further evaluation with larger cohorts and missing modalities.

Closed-form diagonal Gaussian fusion keeps aggregation inexpensive, requiring *O*(| *S* | ∑_*l*_*dl*) arithmetic across fusion levels. End-to-end cost also depends on the encoders and decoders, as reflected by the parameter counts and GFLOPs in Appendix C. The diagonal formulation does not capture general latent correlations, and global weights cannot respond to subject-specific modality quality. Future work could examine subject-dependent weighting, richer covariance models, and the separate effects of decoder structure and modality embeddings.

## 6. Conclusion

We presented task-adaptive Wasserstein barycentric fusion for multimodal medical imaging, which learns subset-normalized modality weights through the application objective and decodes multilevel shared representations using modality-specific decoders conditioned on learnable embeddings. The proposed *Wℬ* VAE-TA combines global fusion weights learned through the application objective with multilevel shared stochastic representations and deterministically conditioned modality-specific decoding features. This shared–specific architecture assigns complementary roles to probabilistic fusion and modality-dependent decoding without introducing modality-specific stochastic latent distributions. Across the evaluated tasks, *Wℬ* VAE-TA improved average performance over the competing approaches and the preceding design variants. The segmentation results support its use under complete and incomplete modality combinations, while the normative modeling results provide complementary evidence under a generative objective. These findings support task-adaptive barycentric fusion together with multilevel modality-specific conditioning.

## Data Availability

ADNI data used in this study are publicly available and can be requested following ADNI Data Sharing and Publications Committee guidelines: https://adni.loni.usc.edu/data-samples/access-data/.

## Acknowledgment

This work was supported in part by the National Institutes of Health (NIH) (R01-AG067103). Computations were performed using the facilities of the Washington University Research Computing and Informatics Facility (RCIF). The RCIF has received funding from NIH S10 program grants: 1S10OD025200-01A1 and 1S10OD030477-01.

## Declaration of generative AI use

During the preparation of this manuscript, the authors used GPT-6.1 (OpenAI) to assist with writing the manuscript and improving grammar, formatting, and spelling. The authors take full responsibility for the content of the manuscript.

## Supplementary Material

### A. Properties of Divergence-Induced Barycenters

This section makes explicit how the orientation and geometry of a divergence determine the fused distribution. Let *S* be a set of observed modalities, let *q*_*m*_ denote the encoding density for modality *m* ∈ *S*, and let 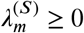 with 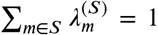. To avoid ambiguity in the terms forward and reverse KL, we write the arguments of each divergence explicitly.

#### A.1. KL Barycenters

##### Component-to-barycenter KL

Define the arithmetic pool 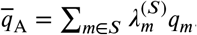. For any normalized density *q*,

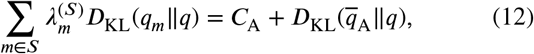

where *C*_A_ is independent of *q*. Hence,

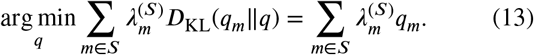

The optimizer is the arithmetic mixture used by MoE. Its support is the union of the component supports, so probability mass retained by any positively weighted component remains represented in the fused density.

##### Barycenter-to-component KL

Let

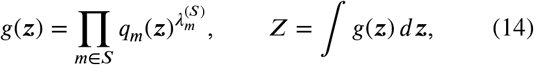

and assume 0 < *Z* < ∞. Then

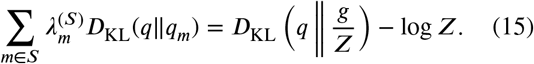

Therefore,

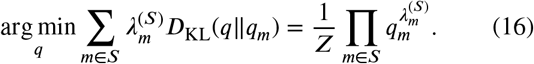

This result is a normalized weighted geometric pool. The standard PoE instead uses unit exponents, giving a precision-sum product for Gaussian components. With positive weights, it assigns zero density wherever any component has zero density; its support is therefore contained in the intersection of the component supports. These identities recover the PoE and MoE relationships used in the barycentric view [12].

### A.2. Gaussian Specializations

For Gaussian components *q*_*m*_ = *N* (***µ***_*m*_, Σ_*m*_), the geometric pool in Eq. (16) is Gaussian. Writing 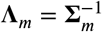, its parameters are

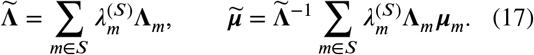

Thus, KL geometric pooling averages Gaussian natural parameters. By contrast, the arithmetic pool in Eq. (13) is a Gaussian mixture and is generally not a single Gaussian. Its mean is 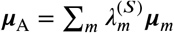, and its covariance includes both within-component and between-component variation:

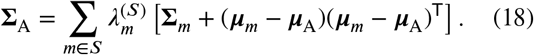

The squared 2-Wasserstein barycenter uses ground-space transport rather than density-ratio geometry [33]. For diagonal Gaussian encoders, it remains Gaussian and has

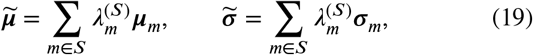

where *σ*_*m*_ contains coordinatewise standard deviations. Consequently, the Wasserstein construction interpolates locations and scales directly, whereas the KL geometric pool averages precisions and precision-weighted means. For general covariance matrices, the Gaussian Wasserstein barycenter is characterized by the fixed-point equation in Eq. (4); the diagonal case yields the closed form used in this work.

### A.3. Sensitivity of Learned Fusion Weights

The following derivation applies separately at each fusion depth *l*; we suppress *l* on the logits, weights, and Gaussian parameters. For *j* ∈ *S*, let the subset-normalized weights be

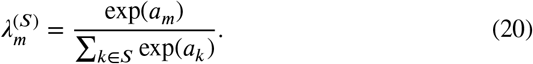

The softmax derivative is 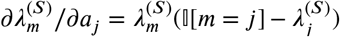. Combining this identity with Eq. (19) gives

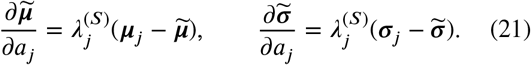

Thus, the local effect of a modality logit is proportional to its current weight and to the displacement of that modality’s parameters from the current barycenter. For *j* ∉ *S*, the derivative is zero because the absent modality is excluded from the subset normalization. This also shows that singleton fusion is insensitive to the logits and exactly recovers the available unimodal distribution.

#### A.3.1 Task-Directed Weight Gradients

Fix the encoder and decoder parameters and an observed set *S* at a single depth. Write *v*_*m*_ = (***µ***_*m*_, *σ*_*m*_) and 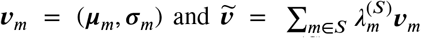 . The softmax derivative gives 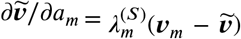. For a differentiable loss depending on the logits only through fusion, the chain rule yields Eq. (8). For the expected loss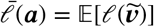, the same identity can be averaged over the training sampling distribution when that distribution is independent of the logits and differentiation and expectation can be interchanged.

A negative derivative in Eq. (8) increases the modality logit under gradient descent, while a positive derivative decreases it. Normalized-weight changes also depend on the other logits at the same depth. Global-weight gradients average these effects across subjects and observed modality subsets. An absent modality has zero derivative for that subset, and a singleton subset provides no weight-learning signal. Each depth has its own logit parameters, so the identity applies to the fused parameters at that depth; gradients are not combined into a shared cross-depth logit. For reparameterized latent samples, the identity applies to differentiable sample losses and their expectations under the stated assumptions.

**Table A.1.**
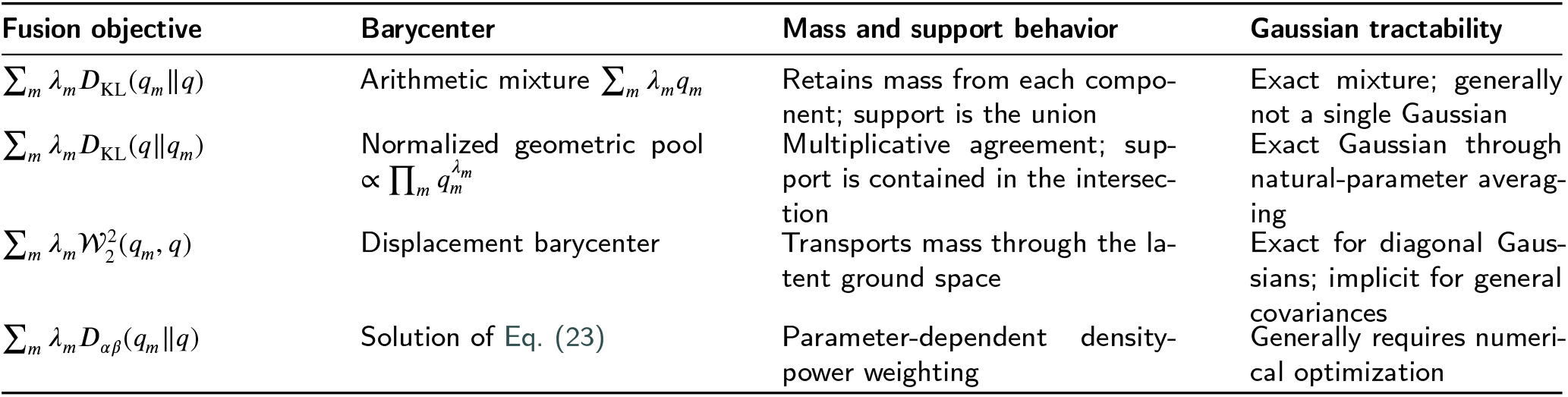
Comparison of divergence-induced fusion operators. KL orientations are written explicitly to avoid naming ambiguity.

**Table B.1.**
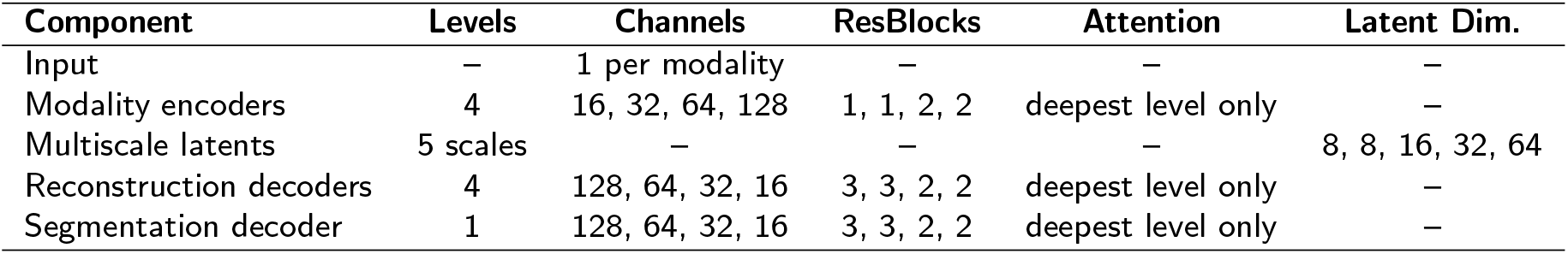
Architecture summary of *W*ℬVAE-TA for brain tumor segmentation.

### A.4. The *αβ* Family and Fusion Geometry

The *αβ* family provides a parameterized connection among density-power divergences [37]. For the definition used in the main text, let

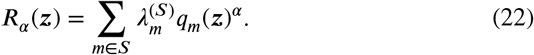

Introducing a Lagrange multiplier η for ∫ *q*(*z*)*dz* = 1, the pointwise stationarity condition for the *αβ*-barycenter is

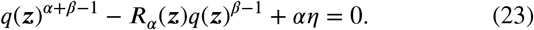

The normalization constant and the nonlinear powers of *q* couple this condition across the latent space, so a normalized closed form is not generally available. The limits (*α, β*) → (0, 1) and (1, 0) recover *D*_KL_(q∥ q_*m*_) and (*D*_KL_q_*m*_∥q), respectively, and therefore recover the geometric and arithmetic pools above. Other parameter choices alter the penalty assigned to mismatched density regions but generally require numerical barycenter estimation.

#### Special cases

For normalized densities, several familiar divergences follow directly from this parameterization:

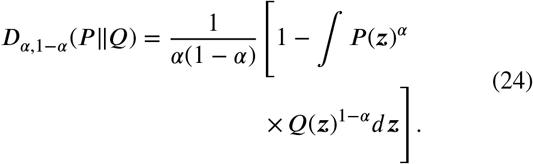

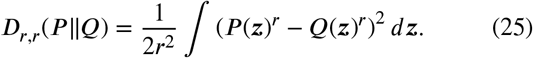

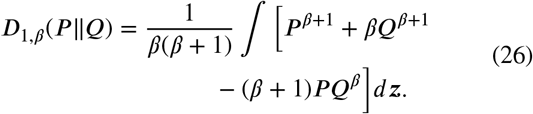

Equation (24), obtained when *α* + *β* = 1, is the alpha-divergence family. At *α* = *β* = 1/2, it becomes 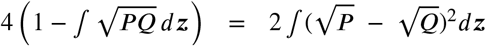, a scaled squared Hellinger distance. Equation (25) shows that *α* = *β* = *r* gives a symmetric power-distance family; *r* = 1 yields 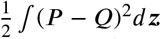. Equation (26), obtained by setting *α* = 1, is the density-power or beta-divergence form under this parameterization. Its limit *β* → 0 is *D*_KL_ (*P* ‖*Q*), it (*α, β*) → (0,1) is *D*_KL_ (*Q*‖*P*). These cases show that changing (*α, β*) alters both symmetry and the relative penalty assigned to low- and high-density mismatches.

#### Fusion divergence versus VAE regularization

The divergence *D* in Eq. (2) determines how the unimodal encoding distributions are combined to construct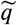. It is distinct from the KL term 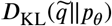 in the ELBO, which regularizes the fused posterior toward the latent prior. Selecting Wasserstein distance for fusion therefore does not replace the variational KL regularizer. These two quantities act at different stages of the model and serve different purposes.

## B. Model Architecture

### B.1. Brain Tumor Segmentation

The *Wℬ* VAE-TA used in brain tumor segmentation is a 3D multimodal VAE. It has four modality-specific encoders, one for each MRI modality: T1w, T1ce, T2w, and FLAIR. Each encoder uses channel widths (16, 32, 64, 128) with residual blocks (1, 1, 2, 2), and applies spatial self-attention only at the deepest 128-channel level. The encoder features are converted into multiscale Gaussian latent variables using 1 × 1 × 1 convolutions. Four latent scales are formed at the encoder levels, and a fifth is formed at the bottleneck. The five latent scales have dimensions (8, 8, 16, 32, 64), which are fused across available modalities by a learnable weighted Gaussian aggregation module. The fused latent features are then mapped back to U-Net feature space and decoded. The model uses four reconstruction decoders, one per modality, plus one segmentation decoder with deep supervision. Each decoder has channel widths (128, 64, 32, 16) and residual blocks (3, 3, 2, 2). Attention is again used only at the deepest 128-channel level, with 16 attention heads. U-HVED [23] uses the same encoder–decoder architecture and configuration as *Wℬ* VAE, *Wℬ* VAE-A, and *Wℬ*VAE-TA, while retaining its original training objective. The detailed architecture is also summarized in Table B.1.

### B.2. Normative Modeling

Since the normative models take preprocessed features as input, we use a simple multilayer perceptron (MLP) architecture for both the encoders and decoders. The MLP architecture has three hidden layers with widths of 32, 64, and 5 units. The latent dimensionality is set to 5. In the normative WB variants, shared latent distributions are formed and fused at all three levels. In the full TA model, a learnable deterministic modality-specific embedding is concatenated with the shared latent sample at the initial decoder stage; subsequent stages combine the preceding decoder features with the shared latent sample at the corresponding level. We use the same MLP backbone architecture for all models reported in Table 4.

## C. Computational Complexity

Table C.1 compares model complexity in the brain tumor segmentation task using an input patch of 128 × 128 × 128 voxels, four modalities, and a batch size of 1. Our model contains 57.62M parameters and requires 220.38 GFLOPs, corresponding to 73.6% and 84.4% fewer FLOPs than DC-Seg and ShaSpec, respectively. Although its parameter count exceeds those of mmFormer and DC-Seg, its computational cost is only 11.2% higher than that of mmFormer. Compared with ShaSpec, our model also reduces the parameter count by 69.3%. The re-implemented U-HVED uses the same encoder– decoder architecture and configuration as our WB-based variants and therefore has similar parameter counts and FLOPs. We report computational complexity for segmentation, which uses a more computationally demanding architecture than the MLP used for normative modeling.

**Table C.1.** Model complexity for an input patch of 128 × 128 × 128 voxels, four modalities, and batch size 1.

| Model | Parameters (M) | GFLOPs |
| --- | --- | --- |
| mmFormer | 37.72 | 198.21 |
| DC-Seg | 40.04 | 833.82 |
| ShaSpec | 187.71 | 1,411.91 |
| Ours | 57.62 | 220.38 |

## D. Learned Modality Weights

For the multimodal segmentation task, modality weights 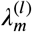 are learned separately at each depth. Their averages across de^*m*^pths are (0.186, 0.358, 0.212, 0.244) for T1w, T1ce, T2w, and FLAIR, respectively. T1ce receives the highest reported weight, consistent with its sensitivity to contrast uptake and its utility in delineating enhancing tumor regions and characterizing the tumor core. FLAIR receives the second-highest weight, aligning with its importance for visualizing peritumoral edema and delineating the whole tumor. T2w provides complementary structural and edema-related information, although its partial overlap with FLAIR may explain its moderate weight. T1w receives the lowest weight, consistent with its primary role in providing anatomical context and its relatively lower discriminative value for tumor subregions compared with T1ce and FLAIR.

As in segmentation, modality weights in normative modeling are learned separately at each fusion level. Their averages across the three levels are 0.46 for T1w MRI and 0.54 for DTI, indicating a nearly balanced allocation between the two modalities. This pattern is compatible with their complementary relevance to AD: T1w-derived measurements characterize gray-matter atrophy, whereas DTI captures white-matter microstructural abnormalities [5]. A similar overall balance is observed in prior multimodal normative modeling work [5]. However, because our weights are learned from healthy individuals under a generative objective, they characterize modality contributions to the normative representation and should not be interpreted as direct measures of AD-specific diagnostic importance.

